# A convergence based assessment of relative differences in age-stratified susceptibility and infectiousness for SARS-CoV-2 variants of B.1.1.7 lineage

**DOI:** 10.1101/2021.03.18.21253931

**Authors:** Sarah D Rasmussen

## Abstract

We propose (a) a method for aggregating and processing age-stratified subregional time series data for positive tests of infection given partial sampling for variant-of-concern biomarkers, and (b) a simple model-based theoretical framework for interpreting these processed data, to assess whether observed heterogeneity in age-specific relative differences can be explained by environmental effects alone.

We then apply this strategy to public-domain subregional time series data with S-gene target failure (SGTF) sampling as a proxy for B.1.1.7 lineage, from weeks 45 to 50 of 2020 from England. For the time period in question, we observe convergence toward a 1.27 (95% CI 1.17-1.38) times higher ratio of SGTF to non-SGTF infection for 0-9-year-olds than for the total population, and a 1.16 (95% CI 1.09-1.23) times higher ratio for 10-19-year-olds. These are roughly comparable to previous findings, but this time we find high-significance evidence for adequate compatibility with our proposed modelling framework criteria to conclude that these relative elevations for 0-19-year-olds are very unlikely to be explained by environmental effects alone. We also find possible indications that 0-19-year-olds might experience a higher relative increase in infectiousness than susceptibility for B.1.1.7.

## Introduction

The emergence of new variants of concern for SARS-CoV-2 brings the challenge of rapidly assessing how a new variant effects transmission, susceptibility, and clinical outcome, relative to the average such effects of previously circulating variants. This includes the more specific challenge of identifying age-heterogeneous effects, and distinguishing such signals of heterogeneity from artefacts of environmental effects.

In late December, a preliminary study by a collaboration of Imperial College and Public Health England researchers^1^ found a statistically significant elevation in relative proportion of infection among 0-9-year-olds (the largest relative increase) and 10-19-year-olds for variants of B.1.1.7 lineage, as detected by sampling for S-gene target failure, which was a strong biomarker for B.1.1.7 lineage in November-Decmber 2020 England.^2^

At the time, various researchers including the present author publicly raised concerns that this observed elevation might be an artefact of environmental factors—specifically increased school transmission during a period of significant restrictions on adult mobility— and that increased data availability and more elaborate evaluation of such data were needed to assess this question adequately.

After a winter school-holiday period involving both a spike in adult cases and a sharp decrease in child and adolescent cases, a Public Health England researcher group performed a new analysis of positive test data averaging in this additional school holiday period, and no longer observed increased relative child and adolescent B.1.1.7 infection in this broader average.^3^ The consensus view then shifted towards regarding prior observations of this signal as having been due exclusively to environmental factors.

In the author’s view, however, neither of these two assessments took adequate account of *time-dependent* behaviour of detected relative infection proportions, which in this case is crucial to detecting potential signals of relative differences in infection unlikely to be explained by environmental effects alone. Meanwhile, increased observations of B.1.1.7-related child outbreaks have accumulated from other countries.^4–6^

Below, we propose a simple method for reprocessing these subregional time series data, and a primitive theoretical framework for interpreting these processed data, based on a model of convergence towards dominant eigenvectors of “transmission” matrices.

## Methods

### Theoretical modelling framework: Background

Since the key strategies of this paper involve convergence towards dominant eigenvectors of transition matrices, we begin by briefly reviewing the notion of a transmission-matrix infection model (a type of non-stochastic transition-matrix model) which is best suited to heterogeneous populations in a relatively constant environment over a time scale with relatively small changes in proportions of population susceptibility.

Restricting to the example of stratifying by age band, let *v*(*t*) denote our infection incidence vector, so that the *i*^*th*^ component *v*_*i*_(*t*) of *v*(*t*) is the number of new infections in age band *i*, per specified time increment ending at time *t*. The model in question then predicts that *v*(*t* + *γ*) = *T* (*t*)*v*(*t*), where *γ* is the generation time of infection and *T* (*t*) is the transmission matrix at time *t*, i.e. the *i, j* entry *τ* (*t*)_*i,j*_ of *T* (*t*) is the expected number of people of age *i* that a person of age *j* will infect. In other words, *τ* (*t*)_*i,j*_ = *c*(*t*)_*i,j*_*p*_*i,j*_, where *c*(*t*)_*i,j*_, the *i, j* entry of a contact matrix *C*(*t*) at time *t*, is the expected number of susceptible contacts of age *i* a person of age *j* will expose, and *p*_*i,j*_ is the probability that an infected person of age *j* will infect a susceptible person of age *i*, given exposure.

If changes in environment or susceptibility proportion are large enough that time dependence of *T* (*t*) has substantial impact, often a more stochastic approach such as a susceptible-infected-recovered model is preferable. When such challenges can be neglected, we take the contact matrix *C*—and therefore the transmission matrix *T* —to be constant in time. Since *T* is ordinarily a positive square matrix, the Perron-Frobenius Theorem then guarantees a unique *dominant eigenvector v*_*T*_ for *T* with all real positive entries and a real positive eigenvalue *λ*_*T*_ with larger absolute value than that of any other eigenvalues. Then for any vector *v* with all positive entries, 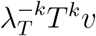 converges to *v*_*T*_ as *k* increases.

Modulo distinctions between infectious period and generation time, this dominant eigen-value *λ*_*T*_ approximates the reproduction number *R*, and this approximation improves as 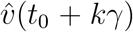 approaches *v*_*T*_. (We use the notation *ŵ* to denote the *L*_1_ unit norm vector *ŵ*:= *w*/| | *w* | |_1_, and normalise all eigenvectors to be *L*_1_ unit norm. Note that if the entries of *w* are non-negative, the *L*_1_ norm is just ||*w*||_1_ =Σ _*i*_ *w*_*i*_).

The above notions are standard. See, eg, work by Munday and colleagues^7^ for recent application of such ideas.

### Key methodological insights

**1. Key theoretical point**. Suppose a variant of concern (VOC) and non-VOC variants circulate in the same environment and elicit robust cross-neutralising activity in convalescent sera (as occurs for variants of B.1.1.7 lineage^3^), so that they share the same contact matrix *C*. Furthermore suppose that changes in environment and susceptibility proportions are small enough for the given time scale to take *C* as approximately constant in time.

Then if the age-stratified infectiousness and susceptibility of the VOC only differ from those of non-VOC variants by an *overall* factor, the respective transmission matrices *T*_VOC_ and *T*_non-VOC_ will differ only by a scalar factor and therefore will have the *same dominant eigenvector*. In particular, if the *L*_1_-unit age-stratified infection incidence vectors 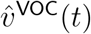 and 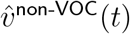 are observed to converge towards significantly different vectors as measured by the limit of the vector 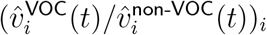 of their entry-wise ratios, then this difference is unlikely to be explained by environmental factors alone.

**2. Key application point**. When aggregating regional time series data for such an analysis, it is helpful to *use VOC proportion of cases as a proxy for time*, or more precisely, to replace time with a synthetic time variable proportional to the logarithm of the ratio of VOC to non-VOC weekly cases in a given region.

To justify point (2), if one aggregates across literal time instead, one risks mixing together regions in different states of VOC spread, which could muddy signals of time-dependent trends including convergence towards dominant eigenvectors. For instance, for any given week, aggregating over literal time risks making regions with earlier VOC spread contribute disproportionately to averages of VOC cases, while regions with later VOC spread contribute disproportionately to averages of non-VOC cases, so that the dominating VOC and non-VOC contributions for any given week have poor comparability. Another advantage of (2) is added flexibility to tune the granularity of time.

To implement point 1, we first assess the appropriateness of the above modelling framework to the data in question, by assessing the convergence behaviour of 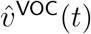 and 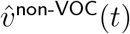, in addition to that of the vector of their component-wise ratios 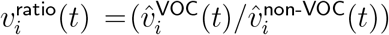. We then estimate the limits to which these vectors converge. The limits of 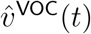 and 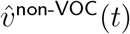 then serve as estimates of the respective dominant eigenvectors (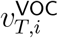 and 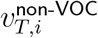)_*i*_. Significant discrepancy of the limit of *v*^ratio^(*t*) from the vector (1, …, 1) is then evidence of age-heterogeneity of VOC effects not caused by environmental factors alone.

However, this *v*^ratio^ does not provide details about changes in relative infectiousness or susceptibility for each age group, or on how the effect sizes of relative infection by age might be impacted by different environments. We therefore construct explicit transmission matrices in the final part of our Results section to explore some of these questions.

### Changes in susceptibility proportions

We pause here for a brief technical point on decreasing proportions of susceptibility, to point out that this is less of a concern in the current case of assessing proportions of infection than it would be for estimating absolute growth. To be more precise, let *β*_*i*_ denote the proportion of the total population in age band *i*, and for initial conditions at time *t*_0_, suppose a proportion *α* of the population is infected and a proportion *σ*_*i*_ of each *i*^*th*^ age band is still susceptible (i.e. not yet immune).

Using the dominant eigenvalue *λ*_*T*_ of *T* as an approximation for *R* and assuming early convergence to the dominant eigenvector *v*_*T*_, we estimate that the proportion of the subpopulation of age *i* still susceptible at time *t* is roughly 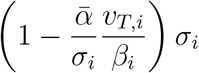, where 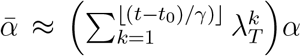. (One could alternatively replace the discrete sum with an integral, if preferred.) For any *i, j* pair, we can then take the cross ratio with the initial susceptibility ratio *σ*_*i*_ / *σ*_*j*_, to obtain 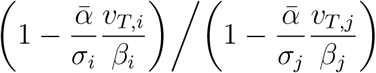.

If these cross ratios are all 1, then changes in susceptibility proportions only effect *C* up to an overall scalar and therefore do not impact dominant eigenvectors or convergence towards them. In the case at hand, we had *σ*_*i*_ ≳ 85% and 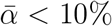, so the effect should be relatively minor. At most, they might lead to a slight observed decrease in 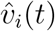 near the end of the time scale for age bands *i* with larger 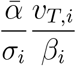, such as for 10-19-year-olds, who had the largest such coefficient. Similarly we would expect the opposite effect in the opposite circumstance. In the actual data, such effects appear to be only slight.

## Applied methods

### Preparation and aggregation of data

In January, the Imperial College team that originally found a disproportionate increase in B.1.1.7 infection in the younger population provided a Github link^1, 8^ with weekly time series data for weeks 43-50 for England, disaggregated into 42 subregions as organised by local health authorities collecting data. The data repository also included population data for subregions and age-stratified population data for England. For each subregion this included a weekly count of positive RT-PCR pillar 2 tests (symptom-targeted community testing)^9^ for SARS-CoV-2, with very roughly 20-35 % of positive tests coming from a “TaqPath” Centre that additionally divided positive test data into tests that were positive or negative for S-gene target failure (SGTF) as a proxy for genomic sampling for B.1.1.7 lineage. During a similar time period in England, Public Health England found that SGTF had 99.5% sensitivity for the Δ69-70 mutation associated with B.1.1.7 lineage.^2^

To maximise continuity of environmental factors, we discarded data from weeks 43 and 44 (schools closed for half-term holidays on week 44). Of the remaining 42 *×* 6 subregion-week pairs, we discarded 50 pairs for low TaqPath sample size, with most of these excluding all 6 weeks for a given subregion. These 50 excluded pairs accounted for 14% of positive tests in the data, 6% of which were from the Cumbria subregion. This was a very conservative pruning of data, only excluding a week-subregion pair if there was an age band for which ≤ 4 people were tested for SGTF.

Each remaining subregion-week pair (*r, w*) had 24 data points, 3 for each of 8 10-year age bands capped at under 80 years old. For the *i*^*th*^ age band, these 3 numbers were

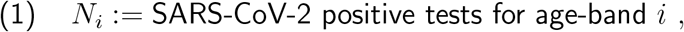

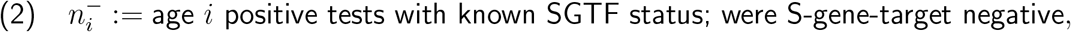

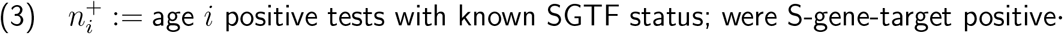

From this, we computed

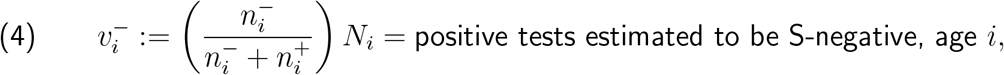

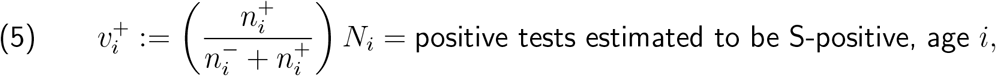

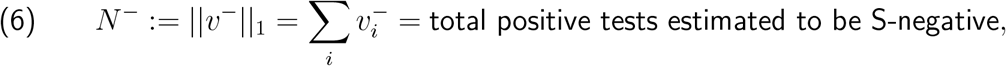

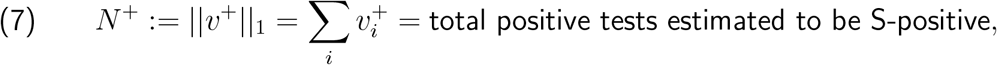

where the − and + labels are respective substitutes for “VOC” and “non-VOC” labels.

For brevity we suppressed (*r, w*) labels in the above notation, but really we should have written 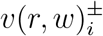, etc. We then convert (*r, w*) to a synthetic time variable, via

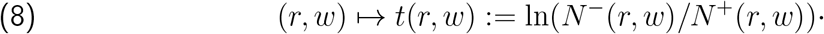

### Smoothing

The next step was to smooth 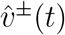 as a function of synthetic time.

For our initial qualitative analysis, we simply grouped synthetic-time-consecutive data into buckets evenly distributed by sample size, as measured by 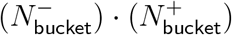. Since 6 weeks is 42 days and the generation time is roughly 5 days, we started with an 8-bucket smoothing, although since these buckets are grouped by sample size, they are not quite evenly distributed across synthetic time, so not quite spaced at equal generation times apart. Since the data were originally presented at the granularity of 6 time points, we also performed a 6-bucket smoothing.

For quantitative assessments of convergence behaviour, we ignored buckets and used a combination of p-value tests and bootstrap resampling estimates.

### Producing an alternate “Rescaled” version from random-sampled prevalence data

We produced two versions of 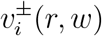 and *v*^ratio^(*r, w*): one from the original, “unrescaled” data, and one “rescaled” version meant to approximate age distribution of infections, as opposed to symptom-based positive tests, by appealing to Office for National Statistics prevalence estimates, which are based on random-sampled RT-PCR nasopharyngeal swab testing of participants taken irrespective of symptom presentation.

Let ***v***^*±*^ := Σ _(*r,w*)_ *v*^*±*^(*r, w*) denote the respective totals of *v*^+^(*r, w*) and *v*^−^(*r, w*) over all subregions and weeks, and let ***o*** denote the vector of average infection-count prevalence per age band for weeks 45-50. To compute ***o***, we took the ONS non-overlapping age-stratified weighted fortnightly estimates for infection-percentage prevalence for weeks 45-46, 47-48, and 49-50,^10^ averaged these percentages for each ONS age band (which were 2-11.5, 11.5-16.5, 16.5-24, 25-34, 35-49, 50-69, 70+), and then, assuming uniform infection rates across ONS age bands, used 5-year age band population data for England^11^ to convert these average prevalence percentages to average infection numbers per 10-year age band, yielding ***o***. Then, with 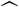 denoting *L*_1_-unit vector, we computed a vector *ω* = (*ω*_*i*_) of rescaling coefficients such that

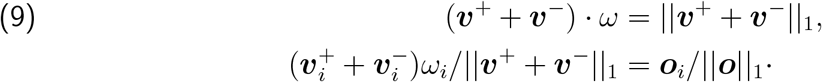

Then to rescale the actual data, we peformed the maps

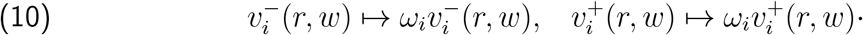

The distinction between symptomatic case count versus infection count is not a pedantic one, when studying age-stratified data. In particular, symptom-based testing in England has typically undercounted 0-9-year-olds by a factor of roughly 2.5 more than the total population, compared to random-sampled prevalence data. Our computed rescaling coefficient for 0-9-year-olds for the relevant time period was 2.42.

On the other hand, it is likely that average infectiousness is lower among the total group of infected persons than among those discovered from symptom-based testing. As such, it is worthwhile to study both the unrescaled (symptomatic cases) and rescaled (estimated infections) data sets in tandem, at least for purposes of studying the vectors *v*^−^ and *v*^+^.

For the ratio *v*^ratio^ itself, the effect of rescaling mostly cancels out, and any observable differences are due to small differences in sorting of data by SGTF proportion, mostly caused by a few small-SGTF-testing-sample-size outliers. Rescaling otherwise only multiplies *v*^ratio^ by an overall scalar very close to 1. That is, after convergence, we have

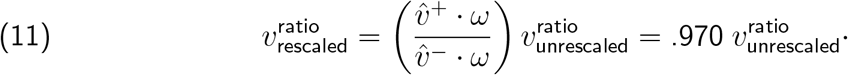

This .970 figure was computed from the quotient 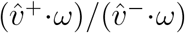 using post-convergence bootstrapped estimates for 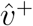 and 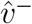. One can use either both unrescaled or both rescaled versions of 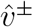 in this computation, as they yield the same answer. For comparison, if one computes 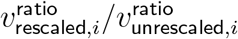 directly from post-convergence bootstrapped estimates, the answers for different *i* range from .969 to .971, so within the limits of rounding error.

This .97 factor simply represents the error inherent in the prior of rescaling *v*^−^ and *v*^+^ the same. With better priors, we could have replaced the rescaling transformation 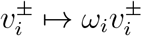 with the transformation 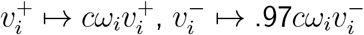, for a scalar *c* close to 1 determined by the equation (*c****v***^+^ + ·97*c****v***^−^) *· ω* = ||***v***^+^ + ***v***^−^||_1_. In particular, since this .97 factor is merely an artefact of minor rescaling error, it is not an intrinsic aspect of 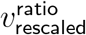, and should be cancelled out.

### Explicit transmission matrix modelling

For the final part of our Results section, we constructed explicit transmission matrix models. To do this, we first started with a contact matrix *C* based on Comix surveillance data from England from the time period in question,^7^ and then constructed an infection probability matrix *P* based on estimates of infectiousness and susceptibilty from data in metastudies, with additional corrections to susceptibility and infectiousness coming from rescaling coefficients, including in the unrescaled case, where, e.g., 0-9-year-old susceptibility must be artificially suppressed to create data consistent with symptomatic testing missing the majority of 0-9-year-old infections. We next adjusted *C* to a matrix *C*^*t*^ so that 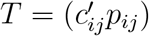 fit the dominant eigenvectors observed. Such *C*^*t*^ is not uniquely determined, so we examined a range of possibilities.

## Results

### (a) Assessment of appropriateness of class of model (assessing convergence)

#### Qualitative assessment of convergence behaviour

We begin by studying graphs of 6-bucket and 8-bucket smoothings of (i) the unit vectors 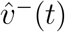 and 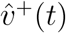 of proportions by age of SGTF (B.1.1.7 proxy) and non-SGTF cases, respectively, as smoothed functions of the synthetic time variable *t* = ln(SGTF*/*non-SGTF); and (ii) the vector *v*^ratio^(*t*) of ratios, 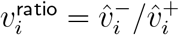. We also examine the versions of these vectors based on non-symptom-based-testing-Rescaled versions of our data. We look for a trend of curves first approaching new values, then approximating horizontal lines.

The graphs of age distribution of SGTF infection (left) and non-SGTF infection (right) in Figure 2 are very nearly a linear transform of the analogous graphs in Figure 1, except that these rescalings cause occasional minor changes in ordering of subregion-week pairs by ln(SGTF*/*non-SGTF), accounting for minor difference in assignments to buckets. The larger separation in curves tends to make the Rescaled data easier to visualise.

**Figure 1.**
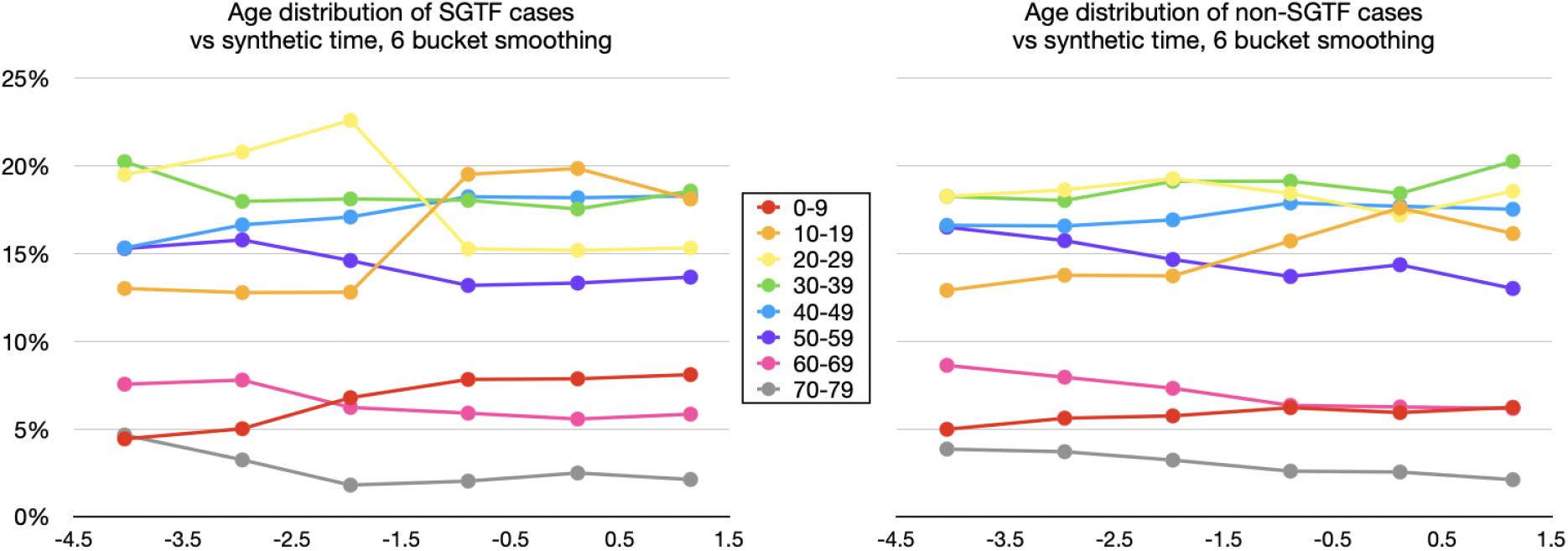
6-bucket smoothings for 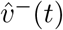—proportion of SGTF (B.1.1.7-proxy) confirmed cases by age-band out of total SGTF confirmed cases—on the left, and for 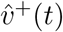 —proportion of non SGTF confirmed cases by age-band out of total confirmed cases—on the right.

**Figure 2.**
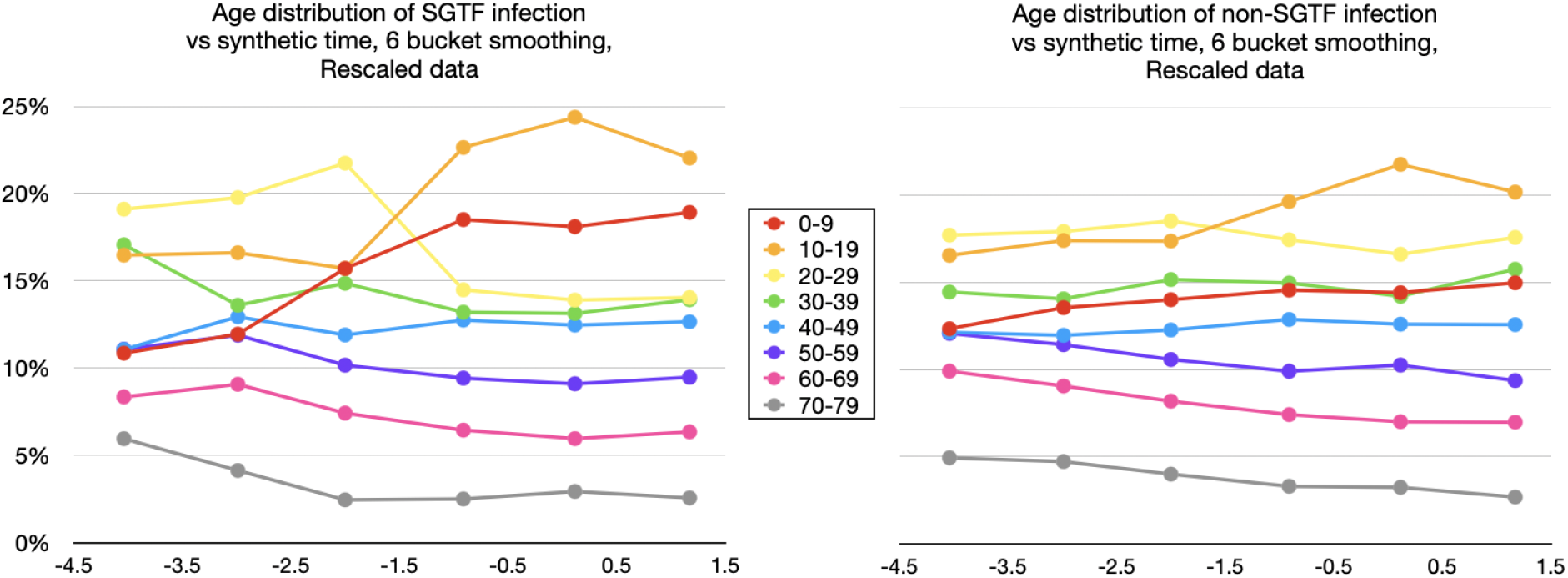
Same as Figure 1, but based on Rescaled data, where positive test data were subjected to a constant overall rescaling by age to match overall age distributions from non-symptom-based testing, to simulate time series data for total infections.

The approach to horizontal lines in graphs of 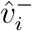 and 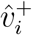 is perhaps easier to see in the 8-bucket smoothings (shown with Rescaled data in Figure 3), due to the greater spread in time. Note how the first 4 synthetic-time buckets of data trend toward a new value, whereas the final 4 buckets tend to hover about a fixed value for each age band.

**Figure 3.**
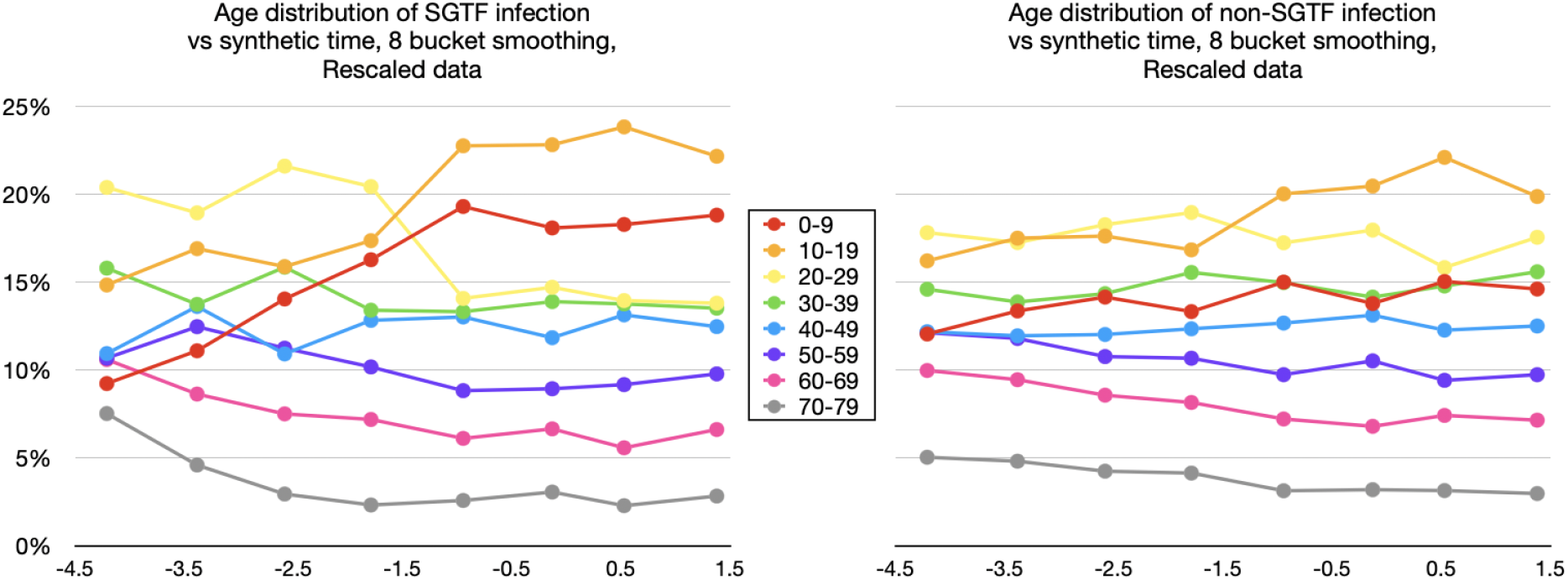
Rescaled version of 8-bucket smoothings for 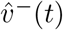 (left) and 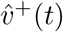 (right).

In Figures 1–3, we see that initial conditions for the B.1.1.7-proxy SGTF graphs (left-hand graphs) are similar to initial conditions for the non-SGTF graphs (right-hand), except with a slight bias towards B.1.1.7 being introduced more by adults. All 6 graphs show an initial relative increase in 0-9-year-old and 10-19-year-old infection. If this seems a surprising trend for the non-SGTF graphs, the reader should keep in mind that the data collection in question started the week immediately following a week-long fall-term school holiday, which coincided with a significant lowering of either infection prevalence or its rate of increase for 0-19-year-olds according to ONS prevalence data, REACT-1 prevalence data, and school absence data.^10, 12, 13^

The right-hand panel of Figure 4 graphs the ratio 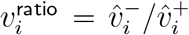 of the two graphs in Figure 3, with the left-hand panel showing the unrescaled version. As noted earlier, the limiting behaviour of the left-hand and right-hand graphs differ from each other by an overall factor of 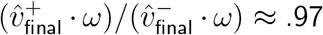, where *ω* is the vector of rescaling coefficients.

**Figure 4.**
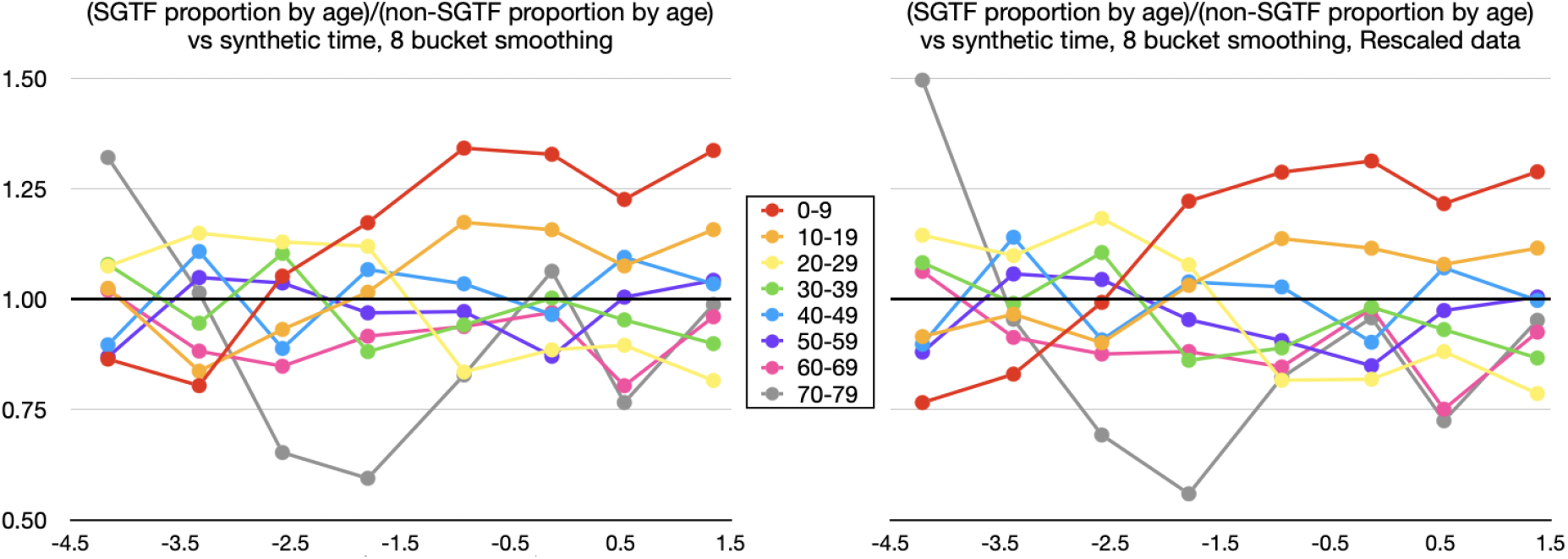
Ratios 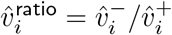, per age band *i*, of corresponding data points from Figure 3 (from Rescaled data) on the right, and from unrescaled data on the left.

The large variation in 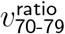 is an artefact of small size of 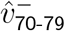 and 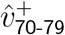, so that ratios greatly exaggerate small variations. Many subregions also had poor sample size for SGTF testing for 70-79-year-olds. Thus, since the final behaviour of the 60-69 and 70-79 bands are similar, we henceforth combine these two age bands.

Although the 6-bucket smoothing for *v*^ratio^ is already smoother than the 8-bucket one, we further improve clarity by graphing fewer age bands at a time in Figure 5. The apparent slight final downward trend for 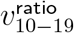 in Figure 5 should be considered in conjunction with the the slight final upward trend for 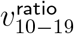 in the 8-bucket smoothing in Figure 8. Note how 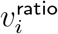 is highest for 0-19-year-olds, but for adults, 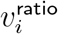 decreases as *i* gets further from 40-49, consistent with Figure 4B by Volz and colleagues.^1^

**Figure 5.**
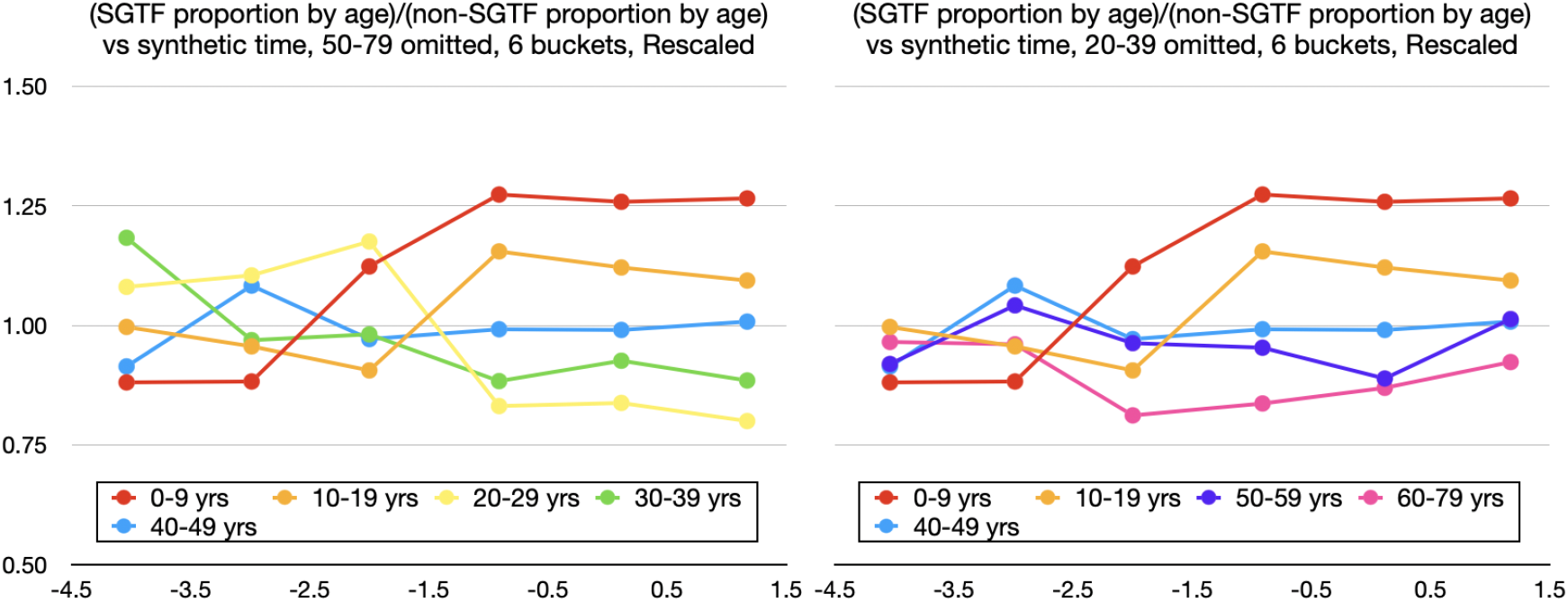
Ratios (*v*^ratio^) of SGTF proportion by age to non-SGTF proportion by age, omitting ages 50-79 on the left, and omitting ages 20-39 on the right. Rescaled version.

We conclude that these data are qualitatively consistent with the behaviour of a transmission matrix model, and that convergence appears to start from approximately the 5th and 4th bucket of the 8-bucket and 6-bucket smoothings, respectively. In each of these cases, the bucket in question started just after a 20% cutoff for total SGTF proportion.

#### Quantitative assessment of convergence

To simplify the signal being measured, we redivide the population into two age bands: 0-19-year-olds and 20-79-year-olds. To produce a prior against which to make comparisons, we use our qualitative assessment cutoff of 20% SGTF to estimate 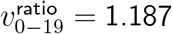 by simply taking the total number of SGTF and non-SGTF cases in each age band for subregion-week pairs with total SGTF proportion *>* 20%, but restricting to TaqPath (SGTF-testing) sample size *>* 20 for each 10-year age-band (with 60-79 combined) per subregion-week.

Returning to our Taqpath sample size *>* 4-per-age-band data, we then conduct 3 series of p-value tests. As a compromise between logarithmic distribution and data availability between cutoffs, we select the cutoffs *>*0%, *>*2%, *>*5%, *>*8%, *>*12%, *>*20%, *>*30%, *>*50%, and *>*70%, for total SGTF proportion for each subregion week. Then, for each cutoff, we conduct

i. a 2-tailed p-value test for number of subregion-weeks with 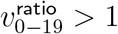,
ii. a 1-tailed p-value test for number of subregion-weeks with 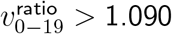, and
iii. a 2-tailed p-value test for number of subregion-weeks with 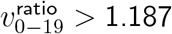.

Test (i) looks for presence of any effect, since no age heterogeneity would imply 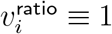. Test (ii) measures when 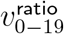 starts trending to be above 1.090—the geometric mean of 1 and our 1.187 prior for 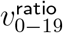. Test (iii) assesses for 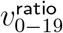 approaching a distribution with median 1.187. Thus, for SGTF proportion cutoffs with ideal convergence, we want low p-values for tests (i) and (ii), but high p-values for test (iii).

To select the best SGTF % cutoff (and hence synthetic time cutoff) for limit estimation, there is a tension between cutting off late enough to have good convergence versus cutting off early enough to have adequate data. In the Table in Figure 6, this tension manifests in a U-shaped pattern for the first two series of p-tests, with minimum at the 20% cutoff.

**Figure 6.**
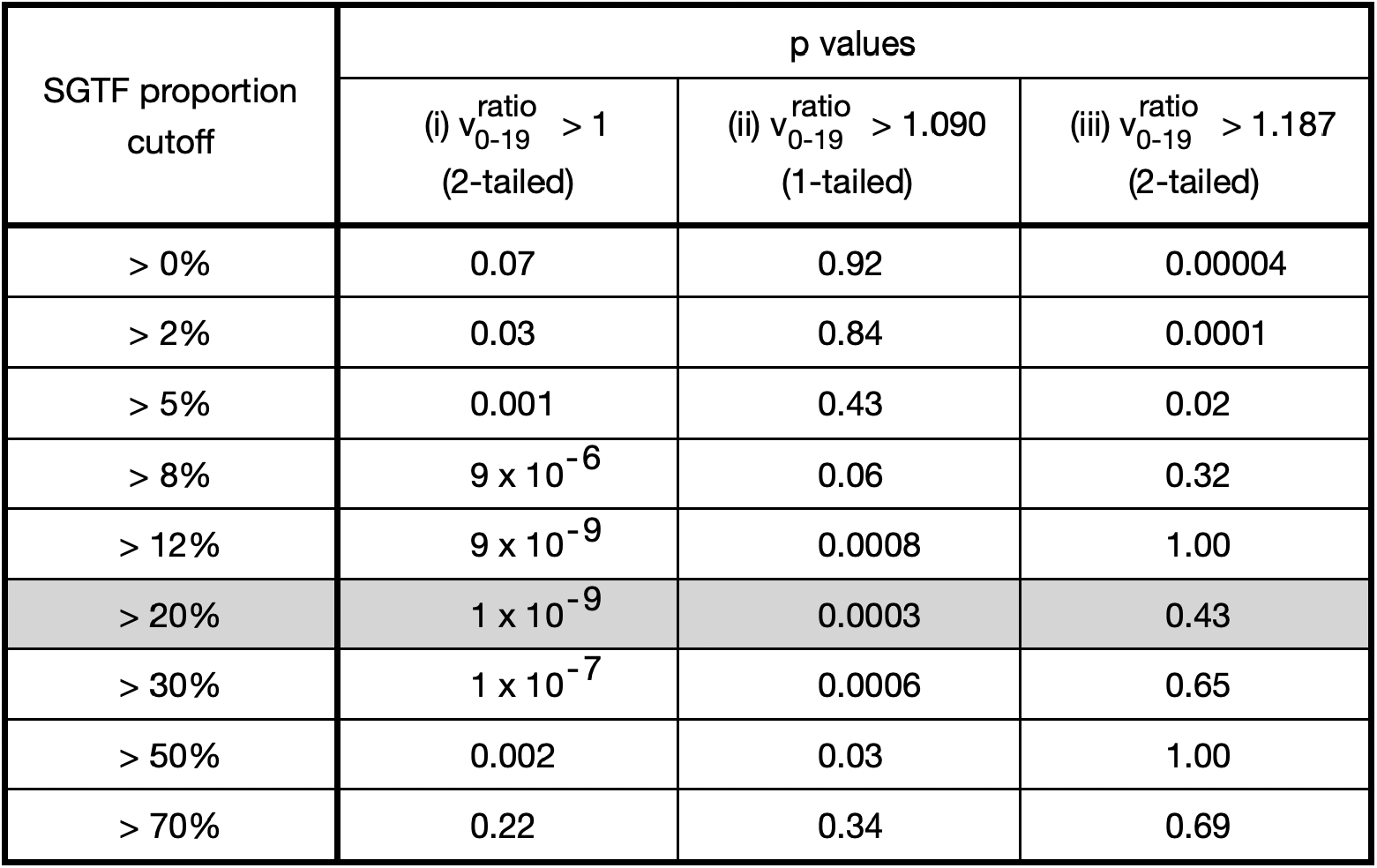
Table of p values to assess convergence at different SGTF proportion cutoffs.

One could worry that sitting at the minimum of the first two series of p tests might make the 20% cutoff biased to have high 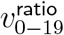, but the Table in Figure 7 shows that is not the case. If one uses bootstrap resampling (which also takes case number into account) to compute *v*^ratio^ point estimates for data from cutoffs higher than 8%, the *v*^ratio^ estimate for the 20% cutoff minimises *L*_2_ distance both from the median and from the geometric mean, and in fact is approximately as close to these two central estimates as these central estimates are to each other. By contrast, the 12% cutoff estimate for *v*^ratio^ overestimates 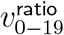, even though it has p = 1 in test (iii).

**Figure 7.**
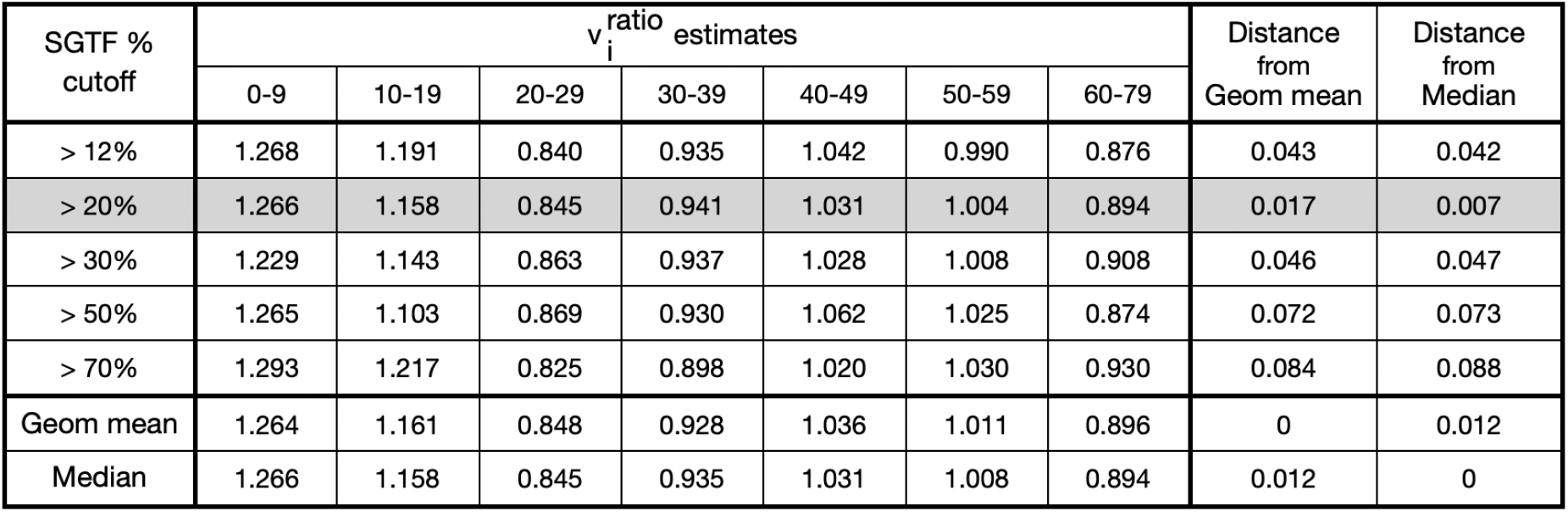
Point estimates for *v*^ratio^ for different SGTF proportion cutoffs, computed from bootstrap resampling, restricted to TaqPath sample size *>*20 per age band.

In summary, we find strong evidence for convergence over (synthetic) time toward a highly statistically significant effect, and we decide on a 20% SGTF proportion cutoff for “post convergent data,” as the shared verdict of qualitative and quantitative methods.

### (b) Estimates of v^ratio^: relative infection ratios for B.1.1.7

Based on the preceding convergence cutoff assessements, we discard all subregion-week pairs with *<* 20% total SGTF proportion, corresponding to the first half of the buckets in the graphs in the preceding subsection. This leaves us with 59 subregion-week pairs. To reduce the role of outliers from small SGTF-testing sample size, we furthermore discard any subregion-week pair with any age band with ≤ 20 cases tested for SGTF (as was also done for the point estimates computed in Figure 8), leaving 50 subregion-week pairs, consisting of about 140,000 positive tests, of which about 75,000 were tested for SGTF.

One could worry these exclusions might exaggerate effect sizes for the 0-9-year-old age band (the source of most low sample-size exlusions), but if anything the opposite is true. These excluded pairs had 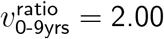 as computed by total cases count, or 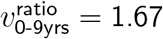 as computed by geometric mean. Seven of these nine excluded subregion-week pairs had a 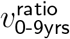 value respectively below or above the respective mininum or maximum of the remaining 50 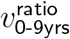 values, and an eighth pair was within 1% of the maximum.

The table and chart in Figures 8 and 9 display point estimates and 95% confidence intervals for *v*^ratio^, computed from bootstrap resampling. (The chart uses slightly higher precision estimates than shown in the table.) Note the similarity between Figure 9 and Figure 4B by Volz and colleagues.^1^ The age trends are the same, but our effect sizes for 0-39-year-olds are slightly larger, likely due to our restriction to SGTF percentage *>* 20%.

**Figure 8.**
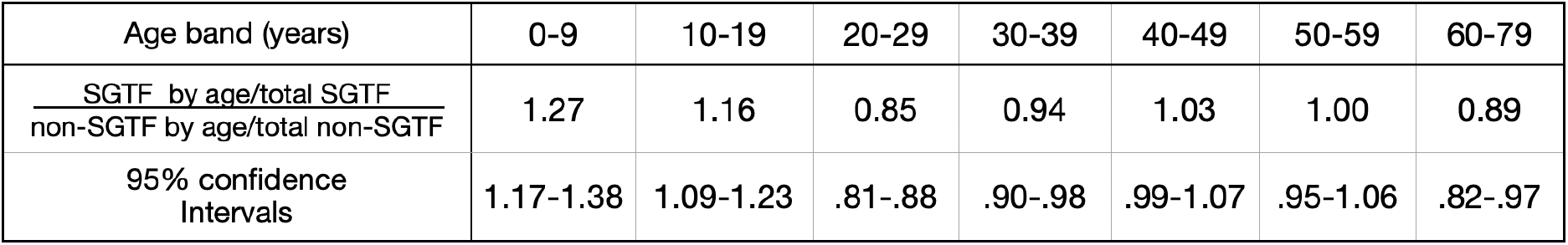
Table of 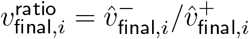 values and 95% confidence intervals.

**Figure 9.**
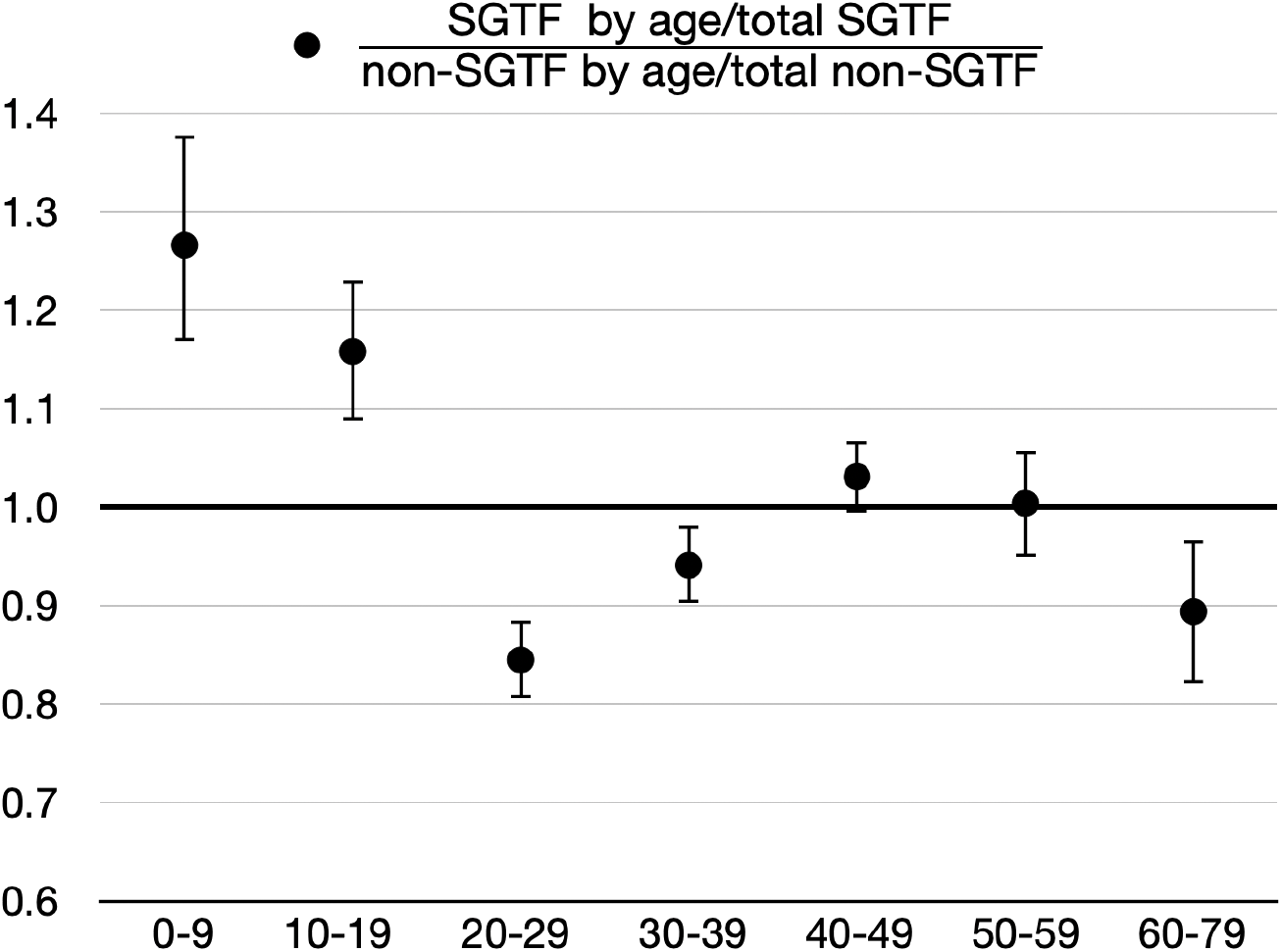
Plot of 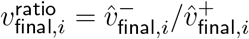 values and 95% error bars.

Again, if the matrix *P* = (*p*_*i,j*_) of probabilities of an infected index in age band *j* infecting a susceptible contact in age band *i*, given exposure, only differed by an overall scalar factor for B.1.1.7, versus non-B.1.1.7 variants, then we should have observed 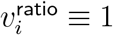.

On the other hand, given that we have demonstrated the *existence* of age-heterogeneous effects in this context, the *sizes* of these effects will be partly influenced by environment, hence the discussion in part (c).

For example, the magnitude of relative infection increase in 0-19-year-olds compared to parent age groups is too large to be explained by an increase in infectiousness in parents, since a relative increase in B.1.1.7 parent infectiousness could only produce that large a relative increase in B.1.1.7 child infection if it produced an even larger increase in B.1.1.7 parent infection. This latter increase in parent B.1.1.7 infection would therefore need to be cancelled out by an extreme reduction in B.1.1.7 parent susceptibility; but decreasing adult susceptibility is equivalent to increasing child susceptibility up to an overall scale.

On the other hand, from fitting matrices *C* and *P* to the available data, there are strong indications that at least part of the inverse U trend for adults in Figure 9 is an inevitable consequence of their gradient of exposure to higher-B.1.1.7-infected 0-19-year-olds.

For illustrative purposes only, we have constructed a very approximate chart of age distributions of parents for Figure 10, to help illustrate the sorts of exposure trends apparent in contact matrices,. To do this, we started with the age distribution of parents of live births in 2017 in England and Wales by 5-year age bands^14^ as the age distribution for parents of a zero-year-old, and used a combination of translations and weighted averages to construct approximations for the age distributions of parents of a primary school student, secondary school student, and 18-year-old, with the idea that among 0-19-year-olds, these groups would be the ones with the largest outside-home exposure.

**Figure 10.**
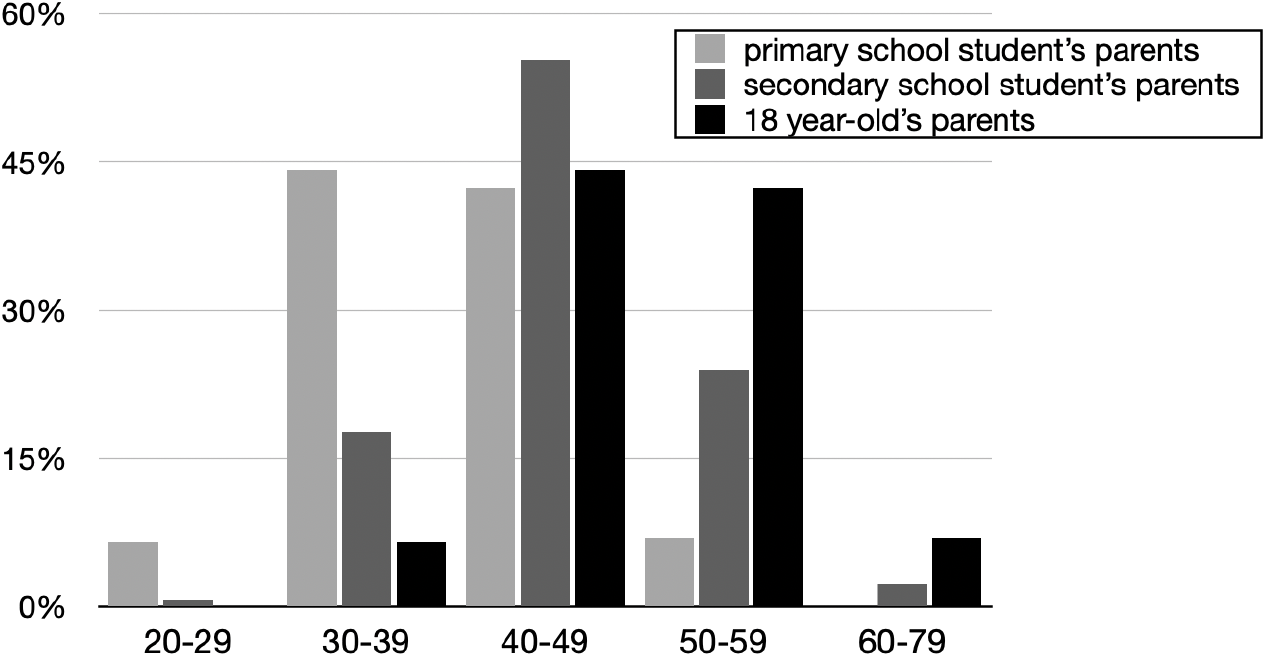
Very approximate age distribution in England and Wales for parents of a person of specified age, in this case a median-aged primary school student, a median-aged secondary school student, and an 18-year-old.

The above analogy between the inverse U trend for adults in Figure 9 and the inverse U trend in exposure in Figure 10 is only qualitative, but such notions can be made precise by contact matrices. We next proceed to direct constructions of transmission matrices.

### (c) Explicit transmission matrix modelling

There are not enough data available in the preceding analyses to determine a transmission matrix *T* uniquely, but one can still construct a range of plausible transmission matrices consistent with the dominant eigenvectors observed.

To improve accuracy at the cost of determinism, we replaced each contact matrix coefficient with a random variable with that coefficient’s expected value. (This change did not significantly impact expected dominant eigenvectors of our matrices.)

We also imposed a temporary damping on *T* at the very beginning of the time series to account for (a) over-dispersion effects from small absolute case numbers early in our SGTF data, and (b) an early-averaging artefact of our synthetic time construction. For (b), when different subregions have their initial weeks introduced at different times early in the synthetic time series, these staggered introductions have a tendency to average out and dampen the transmission progress observed in the first few synthetic time steps. Once most of the subregions are introduced, this damping effect wears off and we observe a progression more typical of the ordinary iteration of transmission matrix multiplication.

In fact, this damping artefact is a powerful asset of our synthetic time construction, as it slows down the convergence process enough to be able to witness it clearly.

In Figure 11, we graph some typical simulation runs for a typical pair of transmission matrices fitting our dominant eigenvector estimates and sharing a common contact matrix.

**Figure 11.**
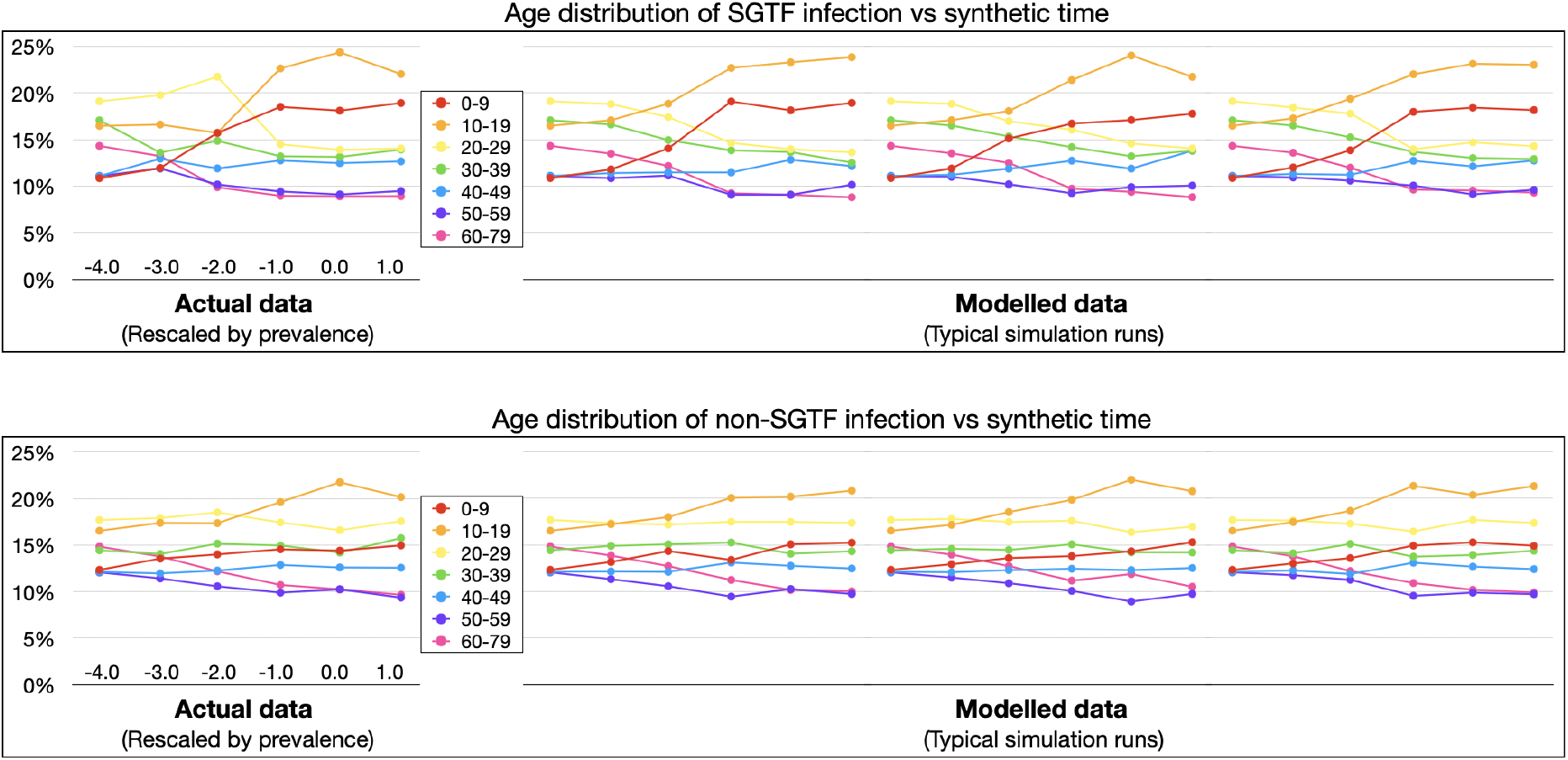
Actual and modelled data for age distribution of SGTF infection (top panel) and age distribution of non-SGTF infection (bottom panel).

To simplify our analysis, we made the assumption that

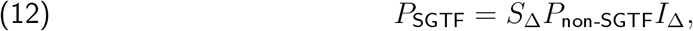

where *S*_Δ_ and *I*_Δ_ are diagonal matrices whose diagonal entries represent relative multiplicative change in susceptibility and infectiousness, respectively, considered only up to an overall scalar factor. (See the beginning of the Methods section for definitions of the contact matrix *C*, infection conditional probability matrix *P*, and transmission matrix *T* = (*c*_*i,j*_*p*_*i,j*_).) This assumption then has the consequence that

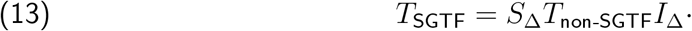

One pattern that quickly emerges is that with any plausible choice of transmission matrix *T*_non-SGTF_ with dominant eigenvector matching 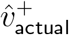, if one increases the infectiousness and susceptibility for 0-19-year-olds and computes 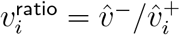 from the new dominant eigenvector 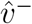, this produces an inverse U trend for adults similar to the trend in Figure 9.

Getting the exact *v*^ratio^ values to match requires tuning the 0-19 values for *I*_Δ_ and *S*_Δ_. In most cases, a better match is obtained from implementing a higher increase in infectiousness than in susceptibility for 0-19-year-olds. Subject to the constraint that 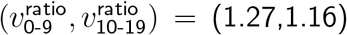, the ratios 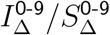 and 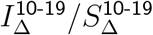 basically control the steepness of the inverse U trend that appears in *v*^ratio^ for adults (using a superscript here to denote diagonal entries of *I*_Δ_ and *S*_Δ_). If these 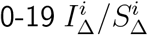 ratios are 1, then the adult inverse U produced is slightly too flat, and one must additionally tune adult infectiousness and susceptibility parameters to obtain a better match.

For example, for one choice of *T*_non-SGTF_, both the pair

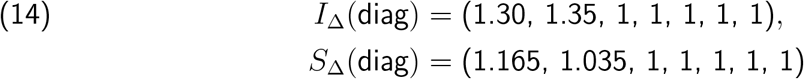

and the pair

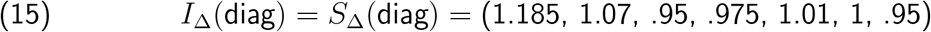

produce *v*^ratio^ with *L*_2_ distance *<* .01 from 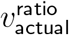, whereas if we demand that 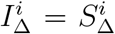 and that all adults have 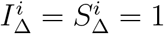, the best we can do is

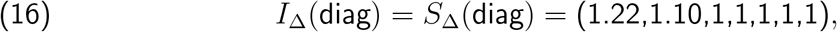

which produces a *v*^ratio^ with *L*_2_ distance .06 from 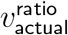. (Note again that *I*_Δ_ and *S*_Δ_ are only defined up to an overall scale. Thus, although we normalised at 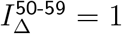, if we divide (15) by the mean for adults, we obtain 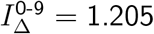.)

If we demand 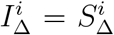 and normalise so that the arithmetic mean of 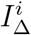 for adults is 1, then for plausible fitting 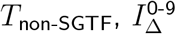 tends to vary from 1.18 to 1.23 as a subjective observation. To produce an objective confidence interval would require systematising what we mean by “plausible” for *T*_non-SGTF_. If 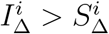, the range of potential values is larger.

#### Sensitivity analysis for other environments

For various fitted choices above, we fixed *P* and (*I*_Δ_, *S*_Δ_) but replaced the contact matrix *C* to reflect a new environmental setting: lockdown with closed schools, or no lockdown and open schools. The original environment was lockdown and open schools, with 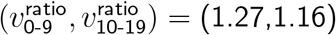.

For a non-systematic selection of choices of fitting *T*_non-SGTF_ and (*I*_Δ_, *S*_Δ_), but with 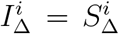, we obtained 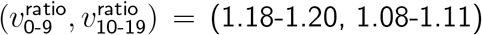 for lockdown and closed schools, and 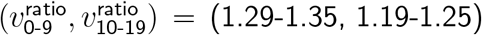 for no lockdown and open schools.

For the same constraints but with 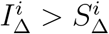 for 0-19-year-olds and 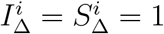 for adults, we obtained 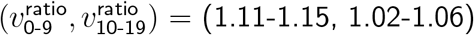 for lockdown and closed schools, and 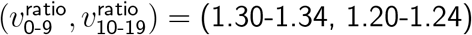 for no lockdown and open schools.

Thus, the age-relative increase of 0-19-year-old B.1.1.7 infection is somewhat more robust to changes in school exposure if 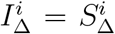 than if 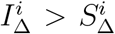, but when the entire community has restored mobility, it matters much less whether 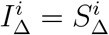 or 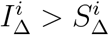.

Again, this was a very non-systematic exploration, and subject to substantial uncertainty about *T*_non-SGTF_.

## Discussion

Before addressing Results obtained, we pause for a warning about analysis methods that risk producing misleading results.

### Comment on averaging accross environments

Subsequent to the Imperial College preprint by Volz and colleagues’ showing increased relative SGTF infection for 0-19-year-olds during weeks 46-51,^1^ the next Public Health England Technical Update on the Variant of Concern included a bar chart in Figure 6^2^ of total symptom-targeted positive tests 1 December 2020 through 4 January 2021 showing a slightly smaller proportion of cases for 0-19-year-olds among SGTF infections than their proportion among non-SGTF SARS-CoV-2 infections.

The Technical Update itself made no comments interpreting this bar chart, but various members of the UK scientific advisory committees SAGE and NERVTAG made prominent comments on social media or media interviews pointing to this bar chart (and similar results) as evidence that the Imperial College results were confounded by environmental factors and that more recent averages showing no B.1.1.7-related relative increase for 0-19-year-olds were a more reliable indicator of relative infection.

Unfortunately, the opposite was true. The Imperial College preprint studied a time of relatively constant environmental factors during which valid conclusions could be drawn about age heterogeneity of relative B.1.1.7 infection. By contrast, the Public Health England chart averaging over cases from 1 December through 4 January mixed incomparable environments in a manner that confounded results.

To demonstrate this latter effect more explicitly, suppose we divide the time interval in question into two intervals: Period 1 from 1 to 20 December, and Period 2 from 21 December to 4 January, with the idea that Period 1 had mostly open schools and more stringent adult restrictions, whereas Period 2 had mostly closed schools and increased holiday-related adult exposure. Let *a*_*j*_ and *c*_*j*_ denote the respective total number of adult cases and child cases in Period *j*, and let 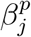 denote the proportion of B.1.1.7 cases in age group *p* and period *j*. There was a sharp decrease in incidence in child and adolescent cases during Period 2, but there was such a peak in adult cases that *a*_2_ *> a*_1_ even though Period 2 was shorter. Thus, regardless of whether “child” refers to 0-9-year-olds or 10-19-year-olds (which we leave ambiguous for the moment), we have *a*_2_*/a*_1_ *> c*_2_*/c*_1_. In particular, if we attempt to compute

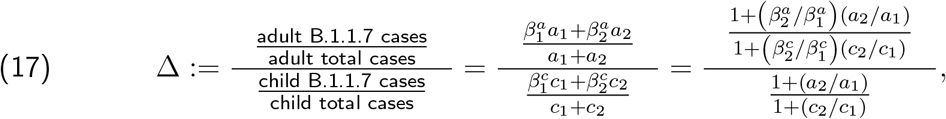

we deduce that even if 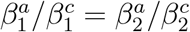, the facts that *a*_2_*/a*_1_ *> c*_2_*/c*_1_ and 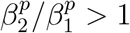 have the automatic consequence that Δ *>* 1.

In other words, the observation that the proportion of 0-19-year-old cases was smaller among SGTF cases than among SGTF cases for the total time period was merely an artefact of the circumstances that many of the child cases occurred when B.1.1.7 proportions were still small, whereas most of the adult cases occurred after B.1.1.7 proportions were larger.

Such averages should not be used to inform notions of the age-stratified patterns of SARS-CoV-2 infection for variants of B.1.1.7, versus non-B.1.1.7, lineage.

### Key Findings

- In an environment of open schools and reduced adult mobility, the ratio (Age-band proportion of SGTF infection)/(Age-band proportion of non-SGTF infection) converged to 1.27 (95% CI 1.17-1.38) for 0-9-year-olds and 1.16 (95% CI 1.09-1.23) for 10-19-year-olds.
- With high statistical confidence, these results cannot be explained by environmental factors alone, and a sensitivity analysis on effect size found that relative elevations in infection for 0-19-year-olds should also be observed in other environments.
- With lower confidence, there are indicators that 0-19-year-olds might experience a higher relative increase in infectiousness than susceptibility for B.1.1.7.

### Potential concerns

Some have questioned the relevance of potential increases in 0-9-year-old infection when this age group is perceived as having such a low infection rate already. This perception, however, is often skewed by the extent to which symptomatic testing undercounts 0-19-year-old infection compared to that of the total population, which for the time period in question was by a factor of 2.4 for 0-9-year-olds and a factor of 1.3 for 10-19-year-olds.

According to symptom-based testing for the time period in question, 10-19-year-olds had a lower confirmed case rate than 20-29, 30-39, and 40-49-year-olds, and 0-9-year-olds had 36% the total confirmed case rate of 20-59-year-olds and the lowest confirmed case rate of any age band.^1^ By contrast, according to random sampled prevalence data, the total infection rate for 20-59-year-olds for that time period was *lower* than that for 0-9-year-olds, and 10-19-year-olds had the highest infection rate of any age band, including more than 50% higher than that for 20-59-year-olds.^10^

One might also be tempted to worry that the age-relative increase in 0-19-year-old positive tests with SGTF merely reflects an age-relative increase in proportion of testing for infected individuals of this age group. For instance, Office for National Statistics questionnaires for participants in their Infection Survey found a slight increase in the proportion of symptoms reported by SGTF-infected versus non-SGTF-infected participants, among those with Ct threshold *<* 30.^15^

However, if the observed relative increase of 0-19-year-olds only reflected an increase in testing rather than an increase in infection, it would be difficult to explain the age trend for relative *adult* B.1.1.7 infection in Figures 8 and 9, which so closely reflects differing exposures to 0-19-year-olds and is consistently reproduced by modelled increases in 0-19-year-old infectiousness and susceptibility. This inverse U age trend for adults was statistically significant: the age 20-29, 30-39, and 40-49 confidence intervals were all pairwise disjoint, as were the age 40-49 and 60-79 confidence intervals.

### Potential mechanisms

One mechanism for our observed age-relative increase in 0-19-year-old B.1.1.7 infection might be the longer viral shedding period proposed for B.1.1.7 in recent findings.^16^ Firstly, there are indications that viral shedding period might be correlated with age,^17^ in which case an increase in viral shedding period might have larger impact on infectiousness of younger age groups. Secondly, an increased viral shedding period would cause a disproportionate increase in transmission in settings of days– or weeks–long exposure. Household transmission is a key example of this, but this would also impact age groups with a higher rate of paucisymptomatic or undetected infection, as subpopulations with undetected infection tend to expose habitual contacts for the duration of their infection.

## Limitations

Whilst it is unlikely that potential disproportionate increases in test-seeking for younger age groups with B.1.1.7 infection could explain observed effects, it is still possible that changes in testing behaviour could impact the precise values of observed effect sizes. It would be useful to see if the Office for National Statistics retain age-stratified SGTF-stratified prevalence data from at least part of the time period in question, as these data are independent of symptom presentation or test-seeking behaviour.

In our “Explicit modelling” subsection, our sensitivity analyses to different environments and to infectiousness/susceptibility mechanisms for observed relative infection increases were limited by high uncertainty about contact matrices, along with mild to moderate uncertainty about infection probability matrices. Even with good surveillance data, accurate contact matrices are difficult to construct, as one must take into account not only the number of people contacted but the duration and quality of exposure. We tried to explore a range of plausible transmission matrices with appropriate dominant eigenvectors, and it appeared that outcomes were not highly sensitive to different choices of matrices, but it is still possible that the range of transmission matrices we explored sat in a qualitatively different component of parameter space from the “true” transmission matrix.

Lastly, while most of our analysis relied on coefficient–wise factorisation of our transmission matrix into separate environmental and intrinsic effects as captured by a contact matrix *C* and infection probability matrix *P*, this approach to some extent exaggerates both the accuracy and the utility of divorcing environmental from intrinsic effects.

For example, there are some indications that a person tends to transmit SARS-CoV-2 more efficiently to a contact of the same age.^18^ Perhaps they speak more to someone of the same age, or perhaps height differentials play a role in close-range droplet and aerosol transmission. If we force our infection probability matrix (or changes to it) to be an exterior product of susceptibility and infectiousness, then we are forced to rely on the environmental contact matrix *C* to capture such aspects of increased transmission efficiency. If certain environmentally-dependent infection patterns are shared by nearly all environmental settings, is this feature best viewed as environmental or intrinsic?

## Data Availability

Data and code for this project will be made available at https://github.com/SarahDRasmussen/b117age.git

## Acknowledgements

The author gratefully acknowledges helpful conversations with Diego Bassani, Deepti Gurdasani, Zoe Hyde, Christina Pagel, and Ashleigh Tuite on aspects of school transmission, testing, increased symptomaticity, and increased viral shedding interval; from Eric Feigl Ding on international B.1.1.7 outbreaks; and from Alex Selby on various statistical methods.

This material is based upon work supported by the National Science Foundation under Grant No. DMS-1926686.

I declare no conflict of interest.

